# Who maintains a good mental health in a locked-down country? A French nationwide online survey of 11,391 participants

**DOI:** 10.1101/2020.05.03.20089755

**Authors:** Frédéric Haesebaert, Julie Haesebaert, Elodie Zante, Nicolas Franck

## Abstract

**Background:** Lockdown measures induce massive societal perturbations and can differentially affect mental wellbeing in populations depending on individual determinants. We aim at investigating the sociodemographic and environmental determinants of wellbeing during global lockdown due to the SARS-CoV-2 pandemic.

**Methods:** A nationwide survey was sent online to the French population during the second week of global lockdown during the SARS-CoV-2 pandemic, between March 25, 2020 and March 30, 2020. Volunteers were recruited on social networks, online newspapers, and mailing lists. We analyzed sociodemographic and environmental variables obtained from a co-built and evidence-based questionnaire. Mental wellbeing was measured by the Warwick-Edinburgh Mental Wellbeing Scale (WEMWBS).

**Results:** We analyzed data from 11,391 (56.6%) out of 20,235 participants who answered the questionnaire. After weighting data on age and gender distributions, 5415 of the respondents were male (47.5%), 5932 were female (52.1%), and 52 (0.5%) registered as “other.” Multivariate analyses indicated that various factors impacted mental wellbeing. Being female (p < .001), a student (p < .001), disabled (p = .001), or having no access to outdoor space (p = 0.02) was associated with lower WEMWBS scores. Conversely, being employed (p < .001) and having more social contacts (p < .01) were both associated with greater mental wellbeing.

**Interpretation:** We revealed differences in mental wellbeing among the French population at the early stages of global lockdown. Authorities should consider the particular vulnerability of students, persons with disabilities, and those living in constrained housing conditions that could increase the negative impact of lockdown on mental health.

## Introduction

Since the initial cases in China’s Hubei province, the coronavirus pandemic (SARS-CoV-2) has progressed to Europe, which became the epicenter in March 2020^1^. To fight virus spread, most countries relied on “old-style” public health measures (e.g. isolation, quarantine, social distancing, community containment)^2^. In line with other countries and informed by models predicting a massive outbreak in the absence of containment measures^3^, the French government enacted a lockdown of its entire population for 15 days, beginning on March 16, 2020^4^.

Massive social restrictions limit face-to-face interactions to the home and during the purchase of necessities. For services deemed non-essential, telecommuting can be enforced, or work can cease entirely. Individuals must reduce their movement to restricted areas that vary depending on living conditions and environment. Thus, lockdown has an immediate and massive impact on daily life. As quarantines and large-scale lockdowns demonstrably impact mental health^5^, we hypothesized that these effects are partially due to social and environmental characteristics. Specifically, changes in routine (e.g., going to work versus unemployment), dissimilar housing types, and variation in social support may generate stratification in mental wellbeing. Early identification of risk factors could help healthcare authorities direct more resources toward vulnerable subpopulations.

With this context, we started a nationwide online survey (LockUwell) during the second week of the lockdown period.

## Methods

We conducted an anonymous cross-sectional online survey in France during March 25–30, 2020. Methodology and reporting of the results are based on the Checklist for Reporting Results of Internet E-Surveys (CHERRIES)^6^.

Participants were recruited with online announcements on social networks, websites of national newspapers, and mailing lists following a convenience non-sampling method, with no incentives.

In line with French regulations on health research, no ethics committee approval was required because data collection was anonymous.

A preliminary version of the “LockUwell” questionnaire was built after gathering information on the lockdown and its psychological effects. The survey included sociodemographic data (section 1), wellbeing from lockdown start (French version of the Warwick-Edinburgh Mental Wellbeing Scale, WEMWBS^7,8^) (section 2), Visual Numerical Scales for stress (section 3), antecedents (section 4), personal situation regarding SARS-CoV-2 (i.e. whether respondents had or knew someone who had SARS-CoV-2 and personal feelings regarding SARS-CoV-2) (section 5), as well as personal and environmental conditions during lockdown (section 6). All sections were presented separately and adapted to specific circumstances. The finalized version was obtained through an iterative testing process that included revisions by researchers, assistants, psychiatrists in several subspecialties, mental-health services users, and citizens. The estimated duration of the questionnaire was 15 to 30 minutes.

Statistics were performed using SAS software 9.4 version (SAS Institute Inc., Cary, NC, USA). The analysis included respondents aged ≥16 years living in France. We weighted data using age and gender distributions from the 2020 French census. In this first brief report, we analyzed data from sections 1, 2, and 6. We described weighted-mean WEMWBS total scores and determinants. Independent variables in the multivariate model were all determinants significantly associated with the total score in bivariate analysis. Multicollinearity was screened using the Variance Inflation Factor (VIF) and the COLLIN option on SAS and no collinearity was found.

## Results

### Descriptive analysis

Of the 20,235 initial participants, 11,742 (58.3%) completed the questionnaire. After excluding respondents with unusable answers and from other countries than France, we ended with 11,391 questionnaires (56.6%) for analysis. After weighting, 47.5% of participants were men, 52.1% were women, and 0.5% were other (**Table 1**). Mean weighted age was 47.47 ± 17.28 years and mean WEMWBS score was 50.51 ± 8.17 (**Table 1**).

**Table 1.**
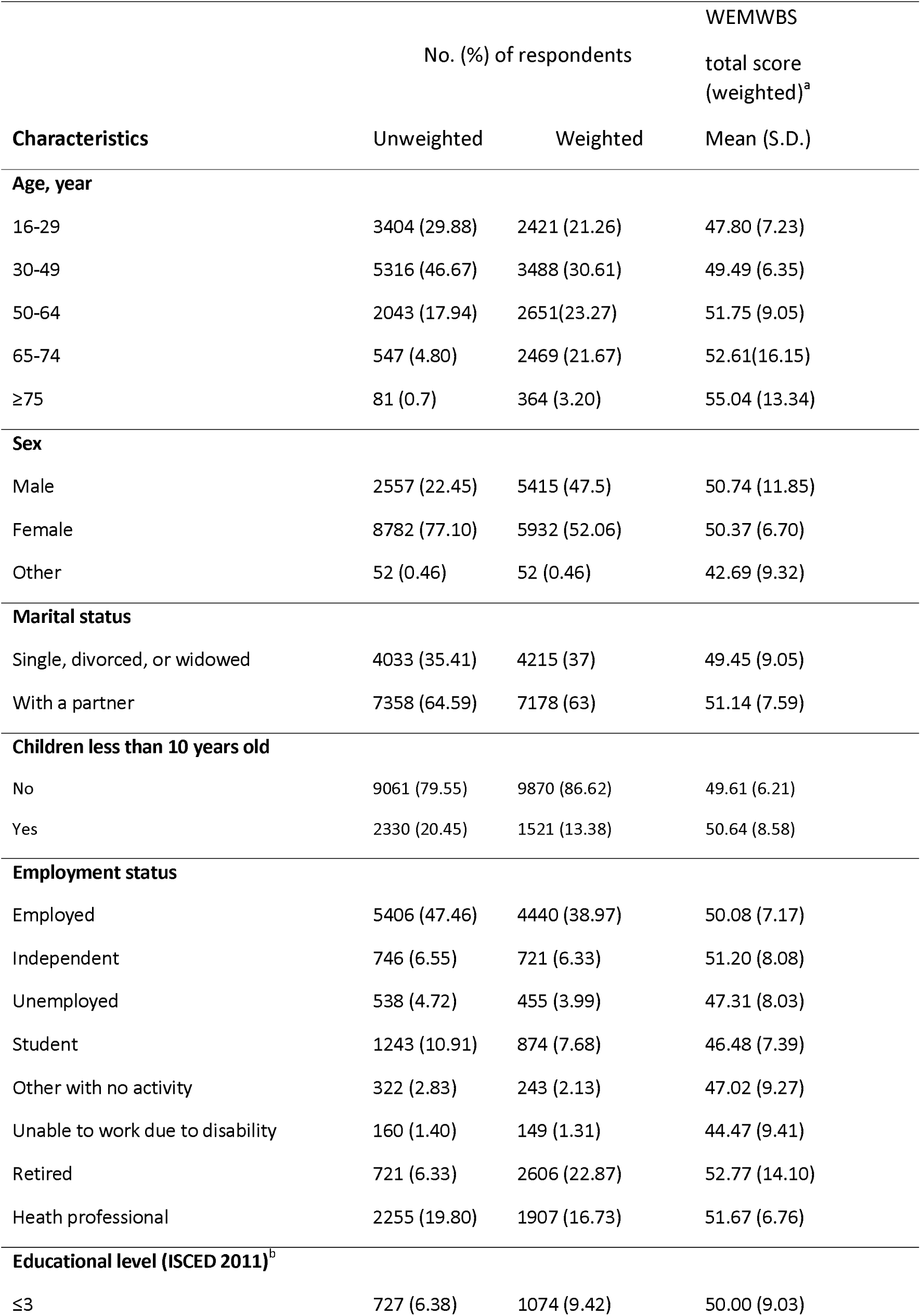
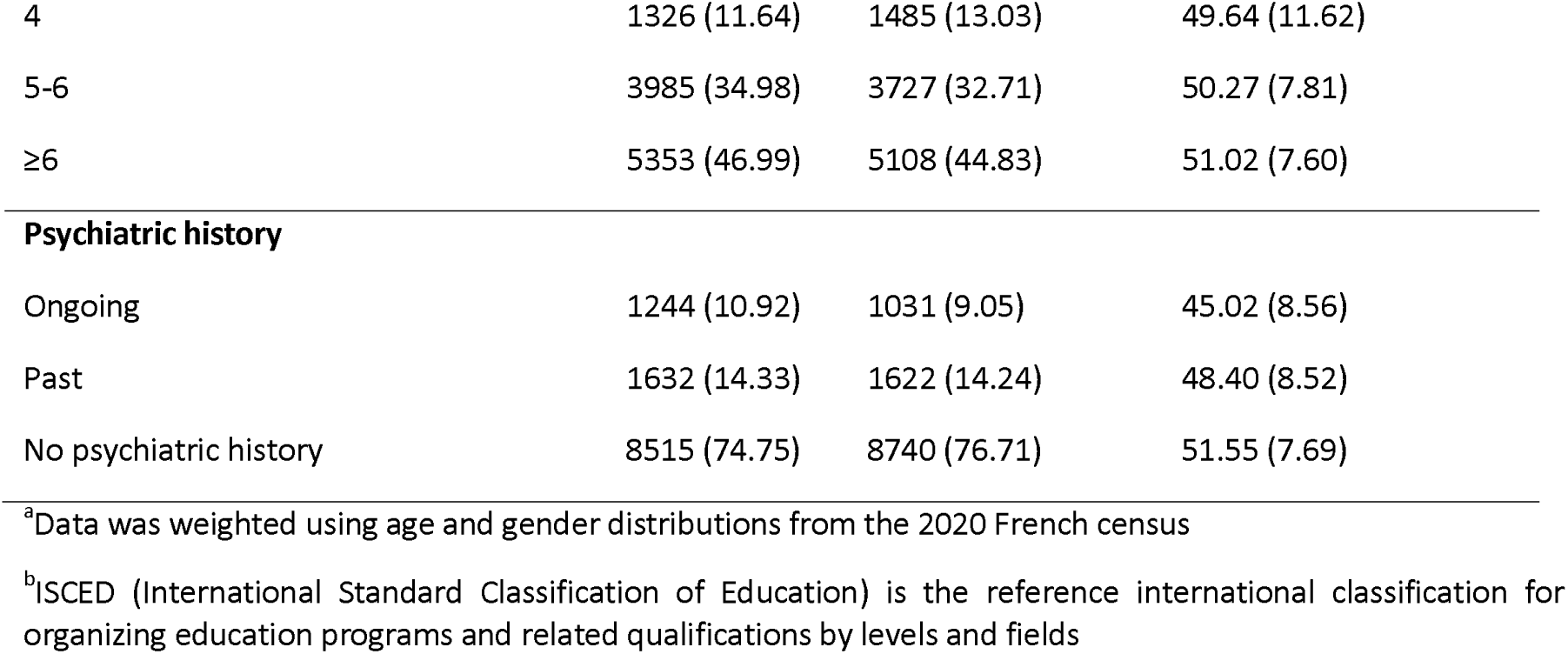
Sociodemographic data of survey participants and their WEWBS total scores (unweighted N = 11391, weighted N = 11393)

| Characteristics | No. (%) of respondents |  | WEMWBS<br>total score<br>(weighted)<sup>a</sup> |
| --- | --- | --- | --- |
|  | Unweighted | Weighted | Mean (S.D.) |
| **Age, year** |  |  |  |
| 16-29 | 3404 (29.88) | 2421 (21.26) | 47.80 (7.23) |
| 30-49 | 5316 (46.67) | 3488 (30.61) | 49.49 (6.35) |
| 50-64 | 2043 (17.94) | 2651 (23.27) | 51.75 (9.05) |
| 65-74 | 547 (4.80) | 2469 (21.67) | 52.61 (16.15) |
| ≥75 | 81 (0.7) | 364 (3.20) | 55.04 (13.34) |
| **Sex** |  |  |  |
| Male | 2557 (22.45) | 5415 (47.5) | 50.74 (11.85) |
| Female | 8782 (77.10) | 5932 (52.06) | 50.37 (6.70) |
| Other | 52 (0.46) | 52 (0.46) | 42.69 (9.32) |
| **Marital status** |  |  |  |
| Single, divorced, or widowed | 4033 (35.41) | 4215 (37) | 49.45 (9.05) |
| With a partner | 7358 (64.59) | 7178 (63) | 51.14 (7.59) |
| **Children less than 10 years old** |  |  |  |
| No | 9061 (79.55) | 9870 (86.62) | 49.61 (6.21) |
| Yes | 2330 (20.45) | 1521 (13.38) | 50.64 (8.58) |
| **Employment status** |  |  |  |
| Employed | 5406 (47.46) | 4440 (38.97) | 50.08 (7.17) |
| Independent | 746 (6.55) | 721 (6.33) | 51.20 (8.08) |
| Unemployed | 538 (4.72) | 455 (3.99) | 47.31 (8.03) |
| Student | 1243 (10.91) | 874 (7.68) | 46.48 (7.39) |
| Other with no activity | 322 (2.83) | 243 (2.13) | 47.02 (9.27) |
| Unable to work due to disability | 160 (1.40) | 149 (1.31) | 44.47 (9.41) |
| Retired | 721 (6.33) | 2606 (22.87) | 52.77 (14.10) |
| Health professional | 2255 (19.80) | 1907 (16.73) | 51.67 (6.76) |
| **Educational level (ISCED 2011)<sup>b</sup>** |  |  |  |
| ≤3 | 727 (6.38) | 1074 (9.42) | 50.00 (9.03) |
| 4 | 1326 (11.64) | 1485 (13.03) | 49.64 (11.62) |
| 5-6 | 3985 (34.98) | 3727 (32.71) | 50.27 (7.81) |
| ≥6 | 5353 (46.99) | 5108 (44.83) | 51.02 (7.60) |
| **Psychiatric history** |  |  |  |
| Ongoing | 1244 (10.92) | 1031 (9.05) | 45.02 (8.56) |
| Past | 1632 (14.33) | 1622 (14.24) | 48.40 (8.52) |
| No psychiatric history | 8515 (74.75) | 8740 (76.71) | 51.55 (7.69) |

<sup>a</sup>Data was weighted using age and gender distributions from the 2020 French census
<sup>b</sup>ISCED (International Standard Classification of Education) is the reference international classification for organizing education programs and related qualifications by levels and fields

### Personal and environmental situation during lockdown

Among the participants, 62.34% had housing with an outdoor space (mean surface area = 75.4 ± 37.2 m^2^). Those living alone comprised 27.73% of participants, while 72.10% lived with at least one other individual. Finally, 15.41% left their homes for work and 33.97% telecommuted. **Table 2** summarizes all lockdown situations and corresponding WEMWBS total scores.

**Table 2.**
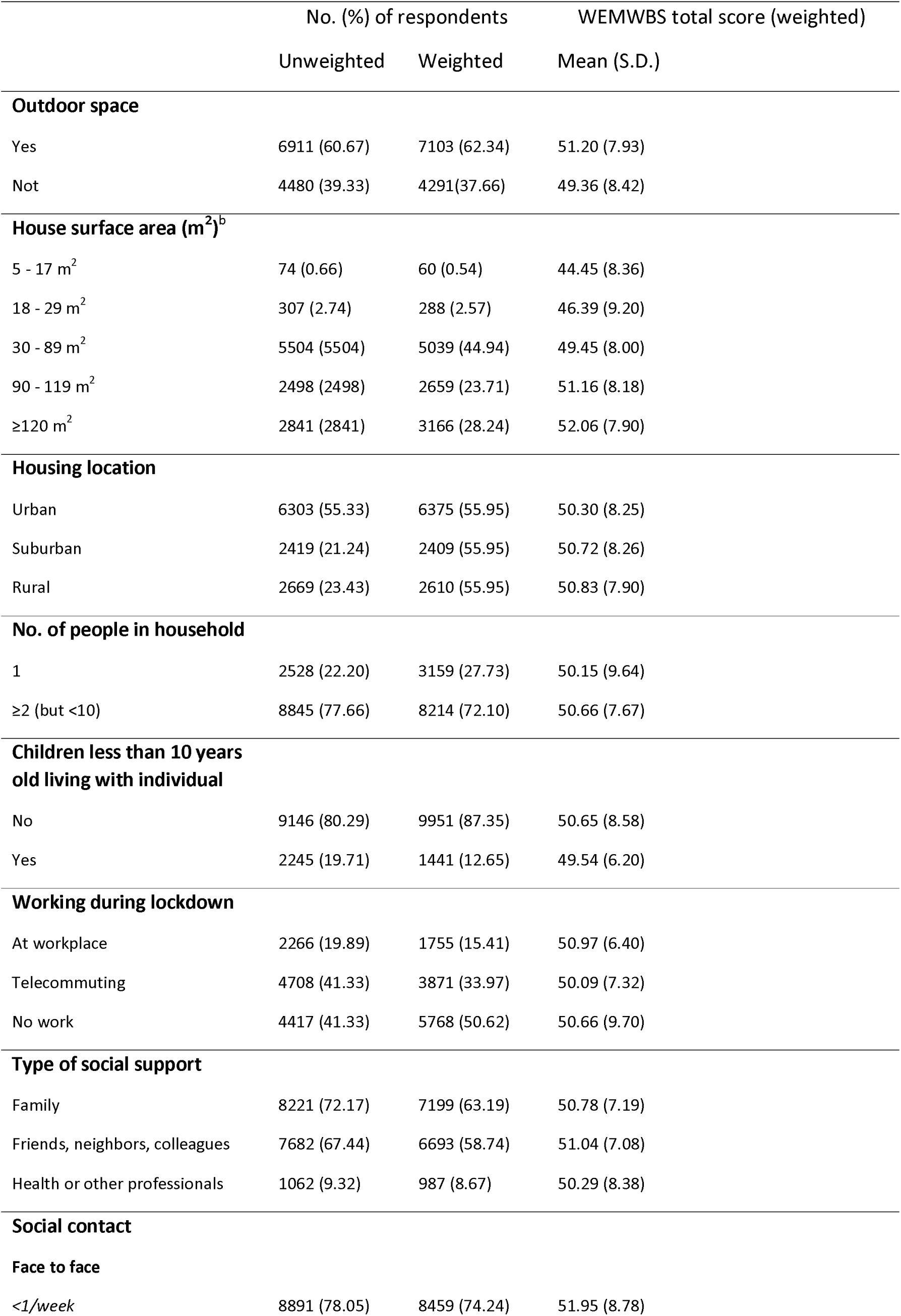
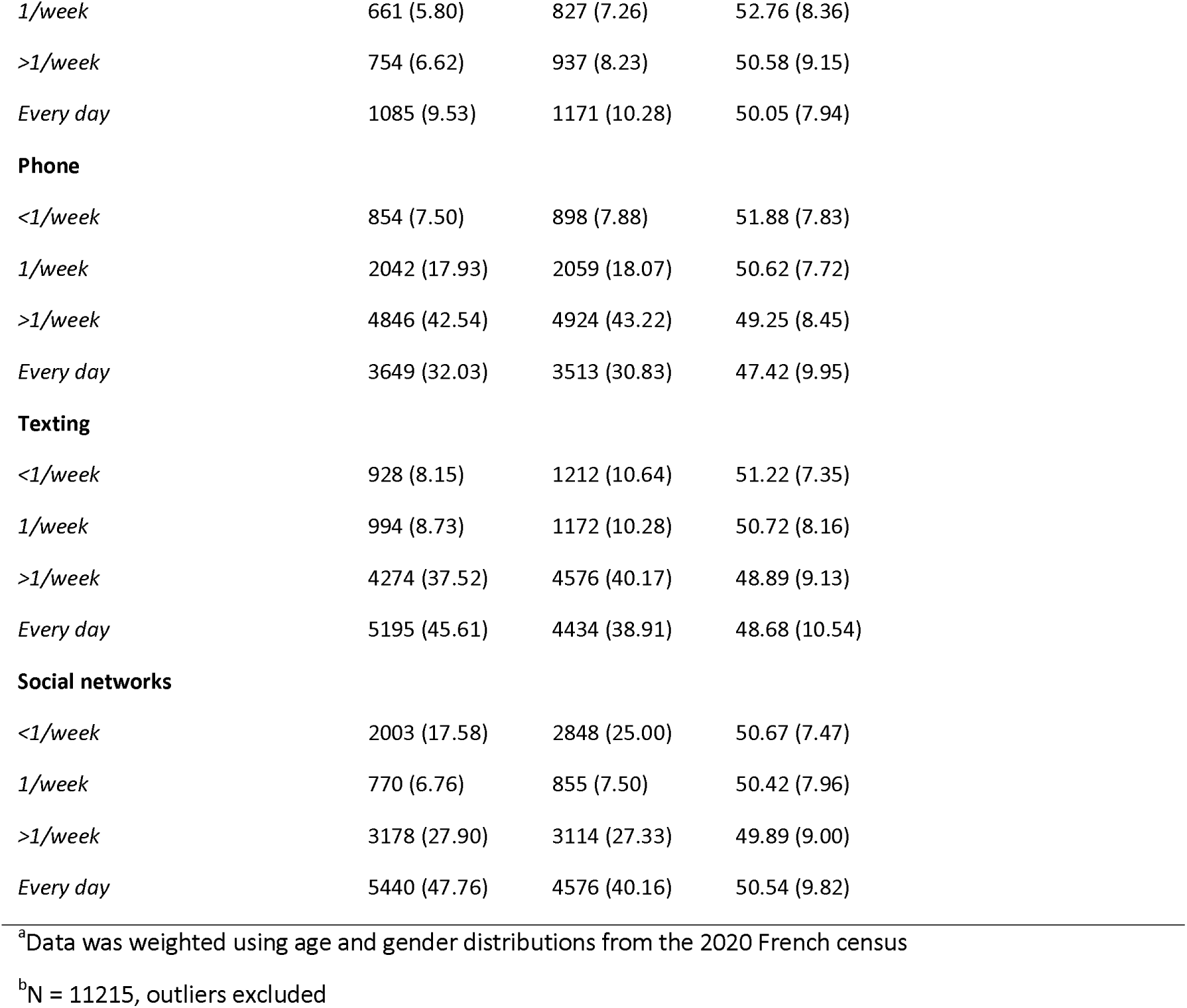
Situation during lockdown (unweighted N = 11391, weighted N = 11393)

### Factors associated with mental wellbeing

Multivariate analyses indicated that being male, having a partner, and being more educated predicted greater wellbeing (**Table 3**). Conversely, having a child under 10 was associated with lower wellbeing. Age was positively correlated with wellbeing. Students and people with disabilities that prevented them from working exhibited the lowest wellbeing scores, whereas retired individuals and healthcare providers had the highest scores. House surface area was positively correlated with wellbeing; in particular, people with access to an outdoor space had higher wellbeing. Wellbeing was also greater among participants who could go to work, had social support, or engaged in higher frequency of social contact via telephone or texting (excluding social media).

**Table 3.** Weighted multiple regression for total WEMWBS scores.

| Determinant | Estimated $\beta$ | p-value |
| --- | --- | --- |
| **Sex** |  |  |
| Other | -3.85 | <.001 |
| Female | -0.95 | <.001 |
| Male | 0.00 | . |
| **Age, year** |  |  |
| 16-29 | -6.46 | <.001 |
| 30-49 | -5.05 | <.001 |
| 50-64 | -2.91 | <.001 |
| 65-74 | -2.21 | <.001 |
| $\geq 75$ | 0.00 | . |
| **Marital status** |  |  |
| Single, divorced, or widowed | -0.51 | 0.007 |
| In a relationship | 0.00 | . |
| **Children less than 10 years old living with individual** |  |  |
| Yes | -0.81 | 0.013 |
| No | 0.00 | . |
| **Employment status** |  |  |
| Independent | 0.56 | 0.062 |
| Health professional | 0.62 | 0.008 |
| Student | -1.74 | <.001 |
| Unable to work due to disability | -2.09 | 0.001 |
| Unemployed | -0.88 | 0.028 |
| Retired | 0.71 | 0.0312 |
| Other with no activity | -1.18 | 0.021 |
| Employed | 0.00 | . |
| **Educational level (ISCED 2011)** |  |  |
| $\geq 6$ | 0.67 | 0.039 |
| 5-6 | 0.01 | 0.982 |
| 4 | 0.00 | . |
| $\leq 3$ | -0.82 | 0.007 |
| **Psychiatric history** |  |  |
| Ongoing | -4.85 | <.001 |
| Past | -2.67 | <.001 |
| No psychiatric history | 0.00 | . |
| **Outdoor space** |  |  |
| No | -0.57 | 0.020 |
| Yes | 0.00 | . |
| **House surface area** |  | <.001 |
| 5 - 17 m<sup>2</sup> | -3.03 | 0.002 |
| 18 - 29 m<sup>2</sup> | -1.23 | 0.013 |
| 30 - 89 m<sup>2</sup> | -0.40 | 0.047 |
| 90 - 119 m<sup>2</sup> | 0.00 |  |
| ≥120 m<sup>2</sup> | 0.76 | <.001 |
| **Housing location** |  |  |
| Suburban | -0.44 | 0.023 |
| Rural | -0.40 | 0.048 |
| Urban | 0.00 | . |
| **Working during lockdown** |  |  |
| No work | -0.94 | <.001 |
| Telecommuting | -0.28 | 0.241 |
| At workplace | 0.00 | . |
| **No. of people in household** |  |  |
| 1 | -0.31 | 0.149 |
| ≥2 (but <10) | 0.00 | . |
| **Type of social support** |  |  |
| **Family** |  |  |
| No | -0.18 | 0.436 |
| Yes | 0.00 | . |
| **Friends, neighbours, colleagues** |  |  |
| No | -1.37 | <.001 |
| Yes | 0.00 | . |
| **Social contact** |  |  |
| Face to face |  |  |
| *Every day* | 0.67 | 0.005 |
| *>1/week* | 1.59 | <.001 |
| *1/week* | -0.36 | 0.187 |
| *<1/week* | 0.00 | . |
| Phone |  |  |
| *Every day* | 1.64 | <.001 |
| *>1/week* | 0.73 | 0.015 |
| *1/week* | 0.31 | 0.319 |
| *<1/week* | 0.00 | . |
| Texting |  |  |
| *Every day* | 1.19 | <.001 |
| *>1/week* | 1.10 | <.001 |
| *1/week* | 0.06 | 0.842 |
| *<1/week* | 0.0 | . |
| Social networks |  |  |
| *Every day* | 1.33 | <.001 |
| *>1/week* | 0.79 | <.001 |
| *1/week* | 0.70 | 0.019 |
| *<1/week* | 0.00 | . |

## Discussion

We report the results of the first nationwide survey on mental wellbeing in a Western European country, at the early stage of global lockdown during the SARS-CoV-2 pandemic. We identified concerning inequities in citizen wellbeing. Notably, students, people with disabilities, and people confined in small spaces with no outdoor access all exhibited lower WEMWBS scores. In contrast, we found greater wellbeing among retired individuals, healthcare professionals, people who could still go to a workplace (instead of telecommuting), and those with more social contacts.

These results are partially in line with other studies on wellbeing during normal (non-pandemic) situations^7,8^, most notably for people with disabilities. However, student WEMWBS scores are far lower than previous research indicates. In a former study, both individual and institutional (i.e., linked to universities) elements could influence student mental health^9^. In the current context, the French student population is faced with cumulative effects from the lockdown: social rupture, closure of universities, and uncertainty about their academic performance^10^. A global lockdown introduces considerable unpredictability for student futures, warranting clear strategies and public messages of hope from the administration and government.

Special consideration should also be given for individuals who live in tiny apartments without outdoor space, especially in urban areas where higher population density makes social distancing difficult, meaning that these inhabitants have few alternatives for maintaining physical activity. Moreover, our results suggest that as lockdowns restrict access to more workplaces, authorities should recommend maintaining social contact via phone and texting.

Finally, as mental wellbeing influences health-related behaviors (e.g., activity and food intake), tracking mental health in the early phase of a global lockdown can inform targeted strategies for improving health and wellbeing among specific subpopulations during and after the pandemic^11^.

### Limitations

This study has some limitations. First, respondents may not be representative of the whole French population due to the sampling method. However, the large sample size and weighting of major sociodemographic characteristics reduce this selection bias. Second, numerous variables influence mental wellbeing, and we are aware that this brief report only included a few of them. Further studies will examine the impact of global lockdown in France and worldwide with greater granularity, but currently there is an urgent need to inform authorities on early determinants affecting mental wellbeing.

## Conclusions

In this French nationwide survey during the second week of global lockdown responding to the SARS-CoV-2 pandemic, we show that mental wellbeing is lower for people with pre-existing vulnerabilities (i.e., disabilities), those who live in environmental conditions that exacerbate social-distancing-related stress (i.e., small spaces), and individuals who are disproportionately affected by uncertainties stemming from the shuttering of institutions (e.g., students). Increased vigilance is warranted in these subpopulations. Policymakers should keep these data in mind when making decisions related to lockdown and post-lockdown strategies.

## Data Availability

No additionnal data available

